# Chlorinated Drinking Water Exposure Enriches Antimicrobial Resistance Pathways in the Infant Gut Microbiome: A Randomized Trial

**DOI:** 10.1101/2024.09.15.24313601

**Authors:** Kimberley Parkin, Claus T. Christophersen, Valerie Verhasselt, Debra J. Palmer, Matthew N. Cooper, Susan L. Prescott, Desiree Silva, David Martino

## Abstract

**Background:** Water chlorination is essential for controlling harmful microbes in drinking water; however, the antimicrobial effects of chlorine-based disinfectants present in tap water may influence early life gut microbial ecology.

**Objective:** To investigate the functional and compositional impact of chlorinated drinking water on the gut microbiome of infants.

**Design:** The waTer qUality and Microbiome Study (TUMS) was an Australian-based double-blinded, randomised controlled trial. Six-month old infants (n=197) received either de-chlorinated drinking water via benchtop filtration (treatment, n=99), or regular chlorinated water (control, n=98) for twelve months. Tap water and stool samples were collected at baseline and at end of intervention. Metagenomic sequencing was used for faecal microbiome analysis. Primary outcomes were differences in gut microbiota alpha and beta diversity. Secondary outcomes included changes in the differential abundance of species-level genome bins (SGBs) and functional profiles.

**Results:** 170 baseline (83 control, 87 intervention), and 130 end of intervention (65 control, 65 intervention) stool samples were collected. Overall community structure was similar between groups after the intervention, including beta diversity (95% CI; 0.4, 0.13, p=0.35), richness (-4.25, 95% CI; -14.85, 6.35, p=0.43) or Shannon Index (0.06, 95% CI; -0.14, -0.32, 0.04, p=0.45). The chlorinated water group showed enrichment of antibiotic resistance MetaCyc groups and pathways (adjusted p < 0.05).

**Conclusion:** Chlorinated drinking water may enhance resistance functions in the infant gut microbiome. While remaining vital for public health, future studies should explore whether adjusting the timing or method of drinking water disinfectants into the infant diet can reduce selective pressures.

**KEY MESSAGES:** *What is already known on this topic:* Chlorination of drinking water is a cornerstone public health intervention to prevent waterborne disease. Chlorine-based disinfectants possess antimicrobial properties that may influence gut microbial ecology, particularly in early life when microbial communities are still developing. Recent studies in low- and middle-income settings have demonstrated that exposure to chlorinated water can increase the abundance of antimicrobial resistance genes (ARGs) in children’s gut microbiota. However, data from high-income settings with advanced water treatment practises, and from randomized controlled trials remain scarce.

*What this study adds:* This randomized controlled trial provides the first evidence from a high-income setting that chlorinated drinking water, while not substantially altering gut microbial diversity in infants, is associated with enrichment of ARG-related functional pathways. The findings indicate that routine water disinfection may exert selective pressures that enhance resistance functions in the early-life gut microbiome, even in the absence of overt taxonomic changes.

*How this study might affect research, practice or policy:* These findings underscore the need to consider AMR implications when evaluating infant feeding guidelines and drinking water disinfection strategies. Further research should investigate whether timing or mode of drinking water introduction can minimize selective pressures and consider alternative disinfection strategies.

## INTRODUCTION

The early life gut microbiota is essential for healthy immune maturation, metabolic development and long-term health ^1^. Perturbations during infancy and early childhood can have long-term consequences for gut health and disease resilience. Antibiotic resistance in children has arisen as a major global health challenge ^2^, with global surveillance studies estimating a 1.8-fold increase in paediatric cases ^3^. The gut microbiota is a potential reservoir for resistance gene acquisition and transmission, and although antibiotic exposure is the primary driver of resistome development, other environmental factors may contribute.

One understudied environmental factor and near universal exposure with the potential to influence antibiotic resistance is chlorinated drinking water ^4^. Chlorine disinfectants are widely implemented and highly effective for controlling enteric infections via the antimicrobial action of chlorine. Current international standards impose an upper limit of 5 mg/L ^5^ on residual chlorine levels in tap water following disinfection, a level considered safe for people of all ages. According to current infant feeding guidelines in Australia, chlorinated tap water can be introduced to infants at twelve months of age, yet there is limited empirical evidence to support this as an optimal time ^6^.

Previous research in animal models has shown the potential for oral chlorine exposure to alter gut microbiome composition, diversity, and function ^7–11^. Observational cohort studies have revealed associations between household water sources and distinct gut microbiota signatures^4,12^. A recent field study conducted in urban Bangladesh linked chlorinated water consumption in children to an increased abundance of ARGs, raising the possibility that chlorination may shape the early-life resistome ^13^. Whether these effects on the gut microbiome are also evident in high-income settings with more advanced water treatment infrastructure and demographically unique microbiota remains an important unanswered question.

The Water Quality and The Microbiome Study (TUMS) was designed to evaluate the effects of residual chlorine on taxonomic and functional changes in the infant gut microbiota. We report results from the first randomized controlled trial (RCT) in which six-month-old infants were assigned to receive either chlorinated tap water or de-chlorinated water for twelve months, and we prospectively evaluated changes in stool microbiota diversity, composition, and function^14^. We report that chlorinated tap water increases the carriage of antibiotic-resistance genes pathways in young children.

## METHODS

### Study design

The Water Quality and the Microbiome Study (TUMS) is a parallel (ratio 1:1), double-blinded randomised controlled trial (RCT), nested within the ORIGINS Project birth cohort ^15^. The ORIGINS Project is a longitudinal observational study of 10,000 pregnant women and their families, recruited from the Joondalup and Wanneroo communities of Western Australia during pregnancy, and followed until their offspring are five years of age ^15^. We recruited six-month-old infants participating in the ORIGINS Project and randomly assigned them to receive either de-chlorinated tap water (intervention group) or regular tap water (control group) for twelve months. De-chlorinated water was provided via a benchtop water filtration device, while the control group received an identical sham filter that did not alter the chemical composition of the water.

Stool samples were collected from participants at study entry (baseline, six months of age) and again after one year (end of intervention, eighteen months of age). Metagenomic sequencing was performed on samples from both timepoints to profile the gut microbiome. Tap water samples were collected at baseline (prior to filter installation, pre-intervention) and post-intervention for detailed chemical analysis. Health assessments were conducted on approximately half of the cohort (opt-in) at twelve months of age as part of the standard ORIGINS Project protocol. The study design is presented in Supplementary Figure 1. The study protocol was approved by the Ramsay Health Care (RHC) WA | SA Human Research Ethics Committee (EC00266; HREC reference number 1911).

### Eligibility criteria

Inclusion criteria were: living within the Perth metropolitan area (Western Australia), born healthy, at least 37 weeks’ gestation, and using tap water connected to the mains water supply to prepare infant drinks. Exclusion criteria were: participating in another diet-based research study, using bottled or filtered water to prepare infant drinks, using childcare facilities for three or more days per week, planning overseas holidays lasting longer than four weeks, having received antibiotics since birth, or being in their seventh month of life.

### Intervention

All participants received a benchtop water filter attached to their kitchen faucet. The filters assigned to the intervention group contained a Hi-Tech Optimizer carbon filtration block rated to remove >97% of chloramines and >99% of chlorine, while preserving fluoride. The control group received identical filters with a sediment-only filter that did not alter the chemical composition of the water. Participants were encouraged to use the provided filter for all infant drinks, maintain normal drinking habits, and carry study-supplied filtered water when outside the home. Filter cartridges were rated for 5,000 litres or twelve months to last the study duration. Participants were advised to follow Australian Infant Feeding Guidelines, regardless of group assignment. Pure Water Systems (Burleigh Heads, Queensland, Australia) supplied the unbranded intervention devices but did not sponsor this study. Both participants and investigators were blinded during the intervention period.

### Randomisation and blinding

Following enrolment, participants were randomised to either treatment group ’A’ or ’B’ using minimization (biased coin; 0.7) stratified by sex, breastfeeding status at recruitment, and mode of delivery. Allocation of filters to treatment groups was done by an independent biostatistician, with both participants and investigators blinded throughout the intervention. The allocator labelled filters, removing identifying information, and revealed group assignments only after data collection was complete. Filters were installed by the principal investigator, and unblinding occurred at completion of the intervention.

### Data collection

#### House visits

Home visits were conducted at study entry and post-intervention by a member of the research team. Baseline data collection included a study entry questionnaire, stool sample collection, tap water sampling, and filter installation. Post-intervention, a water sample was collected from the filter before removal. Participants returned their stool sample kits via a pre-paid envelope.

#### Water quality testing

A sample of flushed (30 seconds) tap water was analysed for heavy metals, major ions, nitrates, pH, phosphates, chloramines, and trihalomethanes at baseline before installing the intervention filter, and at end of intervention. On-site testing for free/total chlorine and pH was also conducted using a Hach D300 pocket colorimeter. Sample collection bottles were filled to exclude air and kept out of direct sunlight. Water samples were delivered to accredited facility ChemCentre in Bentley, Perth.

#### Questionnaires

Participants completed ORIGINS Project questionnaires providing information on antenatal characteristics, water use, child health, breastfeeding, demographics, medical history, and diet. Additional questionnaires assessed filter usage at nine, twelve, and fifteen months. Data was entered into a secure REDCap database.

#### Health assessments

Infant eczema, recurrent wheeze, and allergen sensitization were assessed at twelve months in a subset of participants who opted for paediatrician visits. Medically diagnosed eczema was defined by typical skin lesions clinically observed by a medical practitioner. Recurrent wheeze was defined as more than one separate episode of wheeze symptoms clinically observed by a medical practitioner. Infant allergen sensitisation is defined as a positive skin prick test (with mean weal diameter ≥ 3 mm above the control weal size) to at least one food and/or environmental allergen at one year of age. The panel of allergens tested included hen’s egg, cow’s milk, wheat, fish (tuna), peanut, cashew nut, ryegrass pollen, cat and house dust mite, with histamine and control solutions, using commercial extracts, and in accordance with standard clinical methods^17^.

#### Stool sample collection

Mothers collected stool samples at six and eighteen months of age using provided stool collection kits that utilized a Copan FLOQSwab in an active drying tube (FLOQSwab-ADT, Copan Diagnostics, Murrieta, CA, USA). Samples were shipped directly to Microba Life Sciences Limited (for storage and processing) as previously reported.

### Laboratory processing and bioinformatics profiling

#### Metagenomic sequencing

DNA was extracted from stool samples and subjected to shotgun metagenomic sequencing and microbiome profiling using standard workflows developed by Microba Life Sciences (Brisbane, Queensland, Australia). A more detailed description of the methodology; DNA extraction, library preparation, and sequencing, has been previously published ^18^. To summarise, DNA extraction was performed using a modified protocol, optimising the mechanical lysis step, and following the manufacturer’s instructions for the QIAmp 96 PowerSoil QIAcube HT Kit (Qiagen 47021) on the QIAcube HT automated extraction system. Samples were required to reach a minimum of 0.2 ng/µL to pass quality control requirements. Libraries were prepared according to the manufacturer’s instructions using the Illumina DNA Prep (M) Tagmentation Kit and indexed with IDT for Illumina Nextera DNA Unique Dual Indexes Set A-D (Illumina 20027213-16) with a modification to volume to accommodate processing in a 384-well plate format. Nextera XT libraries were pooled in equimolar amounts to create a sequencing pool and assessed using QuantIT (Thermo Fisher Q33120) and visualised by capillary gel electrophoresis using a QIAxcel DNA High Resolution Kit (Qiagen 929002). Sequencing pools were loaded and sequenced on a NovaSeq6000 (Illumina) according to manufacturer’s instructions and sequenced with 2 x 150bp paired-end chemistry in the Microba accredited laboratories.

#### Raw data processing

Data quality control and processing was performed at Microba Life Sciences Limited. Paired-end DNA sequencing data was demultiplexed and adaptor-trimmed using the Illumina BaseSpace Bcl2fastq2 (v2.20), accepting one mismatch in index sequences. Reads were quality trimmed and residual adaptors were removed using the Trimmomatic v0.39 software package with the following parameters: -phred33 LEADING:3 TRAILING:3 SLIDINGWINDOW:4:15 CROP:100000 HEADCROP:0 MINLEN:100. Human DNA was identified and removed by aligning reads to the human genome reference assembly 38 (GRCh38.p12, GCF_000001405) using bwa-mem v0.7.17 with default parameters, except that the minimum seed length was set to 31 (-k31). Human genome alignments were filtered using SAMtools v1.7 ^21^, with flags -ubh-f1-F2304. Any read pairs in which at least one read mapped to the human genome with >95% identity and >90% of the read length were flagged as human DNA and removed. All samples were randomly subsampled to a standard depth of seven million read pairs to standardize the number of reads across all samples to ensure they could be statistically compared.

#### Taxonomic and metabolic Profiling

Species-level metagenomes were assembled using the Microba Community Profiler (MCP) v1.0 (www.microba.com) using the Microba Genome Database (MGDB) v1.0.3 as the genome reference database. Reads were assigned to genomes within the MGDB, and the relative cellular abundance of species clusters were estimated and reported. Quantification of gene and pathway abundance in the metagenomic samples were performed using the Microba Gene and Pathway Profiler (MGPP) v1.0 against the Microba Genes (MGENES) database v1.0.3. MGPP is a two-step process. In step one, all open reading frames (ORFs) from all genomes in the MGDB were clustered against UniRef90 release 2019/04 using 90% identity over 80% of the read length with MMSeqs2 Release 10-6d92c ^23^. To determine metabolic pathways encoded by metagenome profiles, we used the Enzyme Commission annotations of MetaCyc pathways using the EnrichM tool (https://github.com/geronimp/enrichM), and identified MetaCyc pathways with a minimum completeness >= 80%. In step two, all DNA sequencing read pairs aligned with one or more bases to the gene sequence from any protein within an MGENES protein cluster were summed. The abundance of encoded pathways of species reported as detected by MCP was calculated by averaging the read counts of all genes for each enzyme in that pathway.

### Statistical analyses

All analyses were performed using R version 4.3.1. All boxplots represent the first and third quartiles, with a median as a middle line and whiskers at the last value within a 1.5 × IQR distance from the upper or lower quartile, where IQR is the interquartile range. An alpha level 0.05 was used to define statistical significance for unadjusted and adjusted p-values as appropriate. All participants who successfully submitted a baseline (n= 170) and end of intervention (n=130) were included in the relevant analyses. Participants who did not submit stool samples at the relevant timepoint were not included in that analysis.

#### Sample size

Our sample size was based on a previously published randomised control crossover intervention, which investigated the effects of alkaline and neutral water pH on faecal microbiota in 29 Danish men. The authors noted that their post hoc analysis indicated a sample size of 180 would have 80% power to detect differences alpha diversity at a p-value of 0.05 ^25^.

#### Study outcomes

The primary outcome of this study was the difference in infant gut microbiome composition between the intervention and control groups, measured from stool samples collected at six and eighteen months of age. Secondary outcomes included the incidence of allergic sensitization, wheeze, and atopic eczema, as assessed by physician diagnosis at 12 months of age.

#### Community comparison

The overall composition of the gut microbiome was visualized using taxonomic bar plots of the average relative abundance of the top eight most abundant phyla. Data were stratified by age and intervention group.

#### Diversity analysis

Diversity measures were calculated using the phyloseq R package (version 1.38.0) ^26^. The Shannon index and observed richness were used to estimate alpha diversity and the mean difference between groups was presented with a 95% CI and Student’s T-test comparison. Beta diversity was calculated using the Bray-Curtis dissimilarity index, and non-metric multidimensional scaling (NMDS) was employed to visualize beta diversity. Permutational multivariate analysis of variance (PERMANOVA) was used to formally test for differences in beta diversity between groups ^27^. Beta diversity confidence intervals were calculated using bootstrapping (n=1000).

#### Differential analysis

To identify differentially abundant SGBs and metabolic pathways between the intervention and control groups at 18 months of age, we utilized DESeq2 (version 1.40.2) ^28^. This approach models count data using a negative binomial distribution and applies a Wald test for significance. SGBs present in at least 5% of samples (n=9/170) and with a total count exceeding 1000 were considered for analysis, and taxonomic classification was determined at the species level. P-values were adjusted for multiple comparisons using the Benjamini-Hochberg false discovery rate (FDR) correction. Similarly, differentially abundant MetaCyc pathways between groups were identified using DESeq2. Data visualization was performed using the R packages phyloseq (24), microViz (version 0.10.0) (27), ggplot2 (version 3.4.0) (28), and ggstatsplot (package version 0.12.2) (29).

### SAGER guidelines

Subject sex was recorded in clinical notes as parent-reported male or female. Regression modelling was adjusted for sex such that findings are generalizable to both sexes.

### Inclusion and Ethics

This research was conducted in partnership with local researcher consumers who were involved in the design, data collection, analysis and reporting of the study. The research is locally relevant, and the study protocol was acceptable to parents and local Institutional Review Board (IRB).

## RESULTS

### Study Design and Participant Flow

Recruitment occurred between November 2019 and March 2022, randomising 197 participants to the control (n=98) or treatment (n=99) groups. Baseline characteristics were balanced between groups, including sex, delivery method (assisted vaginal, unassisted vaginal, elective caesarean, or emergency caesarean), and breastfeeding status at recruitment (predominantly formula-fed, predominantly breastfed, or mixed-fed), as shown in Table 1. Further demographic details on the cohort were described previously ^18^. During the study, 76 adverse events (AEs) were reported, defined as instances where a participant presented to hospital for any reason. These were mostly due to unrelated common childhood infections or injuries. Two participants experienced serious adverse events (SAEs) including anaphylaxis, and septic arthritis which were assessed and deemed unrelated to the intervention.

**Table 1.**
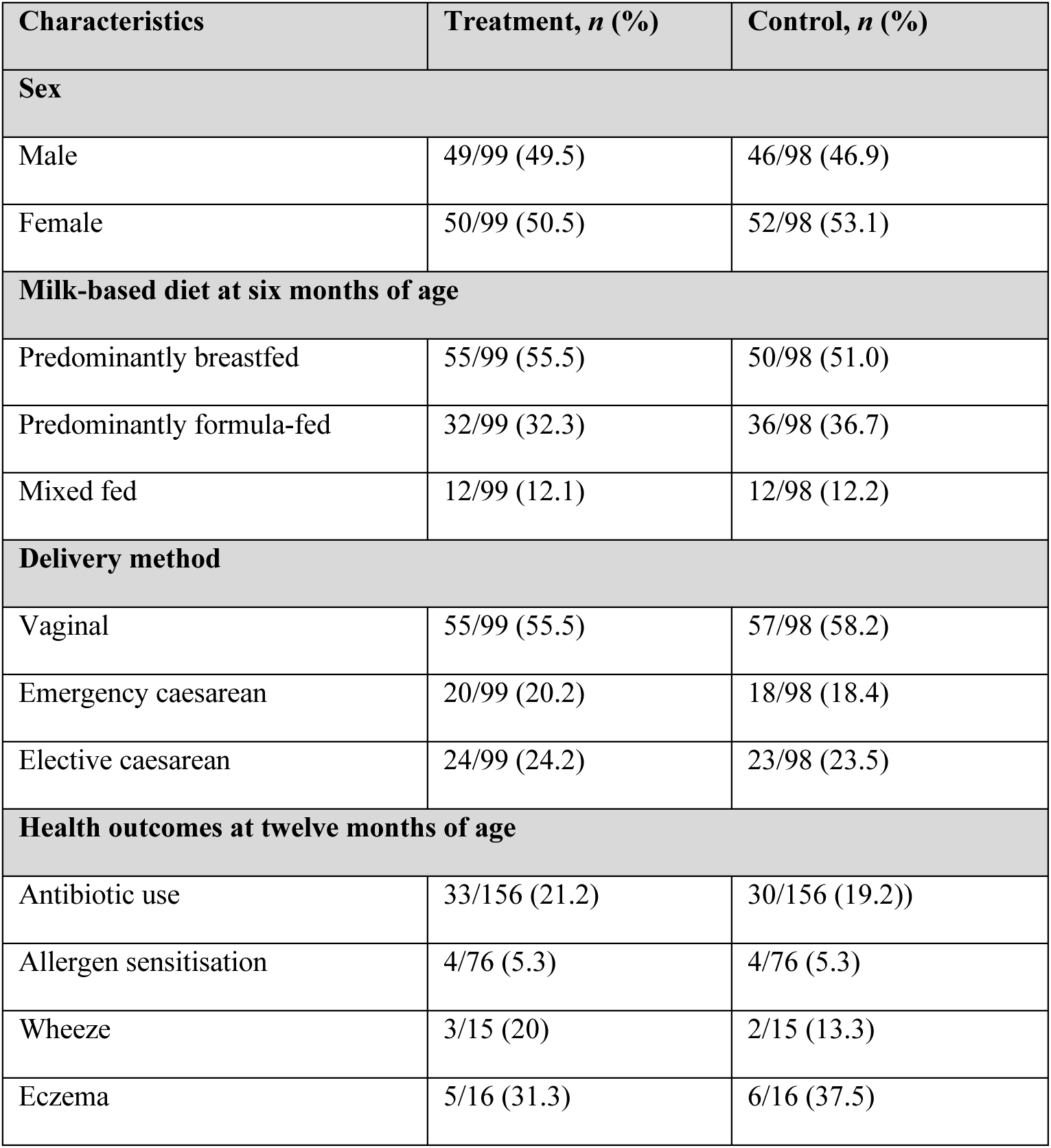
Characteristics of 197 children participating in TUMS.

Of the 426 infants assessed for eligibility, 229 were excluded (Supplementary Figure 2). Reasons for exclusion included: not meeting inclusion criteria (n=190), declining participation (n=36), residing outside the eligible area (n=1), and logistical constraints preventing the initial home visit (n=1). Of the 197 randomised participants, 10 did not receive the intervention (4 treatment, 6 control) due to filter incompatibility with their tap fittings. A total of 170 participants submitted baseline stool samples, and 130 participants (66%) completed the study and provided stool samples at eighteen months of age. Among those with twelve-month health assessment data available, no significant differences were observed in health outcomes between intervention and control groups, including antibiotic use (Table 1). Due to the logistical disruptions related to the COVID-19 pandemic, the rate of missing data in these health outcomes was high (approximately 50%).

### Effectiveness of the Intervention on Water Quality

Participants primarily resided in the northern regions of Perth, Western Australia. Pre- and post-intervention water samples were collected from 83 and 57 treatment group households, and 82 and 57 control group households, respectively. Pre-intervention total chlorine levels varied across homes (0.3-1.1 mg/L), but all were below the 5 mg/L recommended by the Australian Drinking Water Guidelines (Figure 1A). Disinfection by-products (bromoform, bromodichloromethane, chloroform, chlorodibromomethane) also varied at baseline but remained below recommended levels (Supplementary Figure 3).

**Figure 1.**
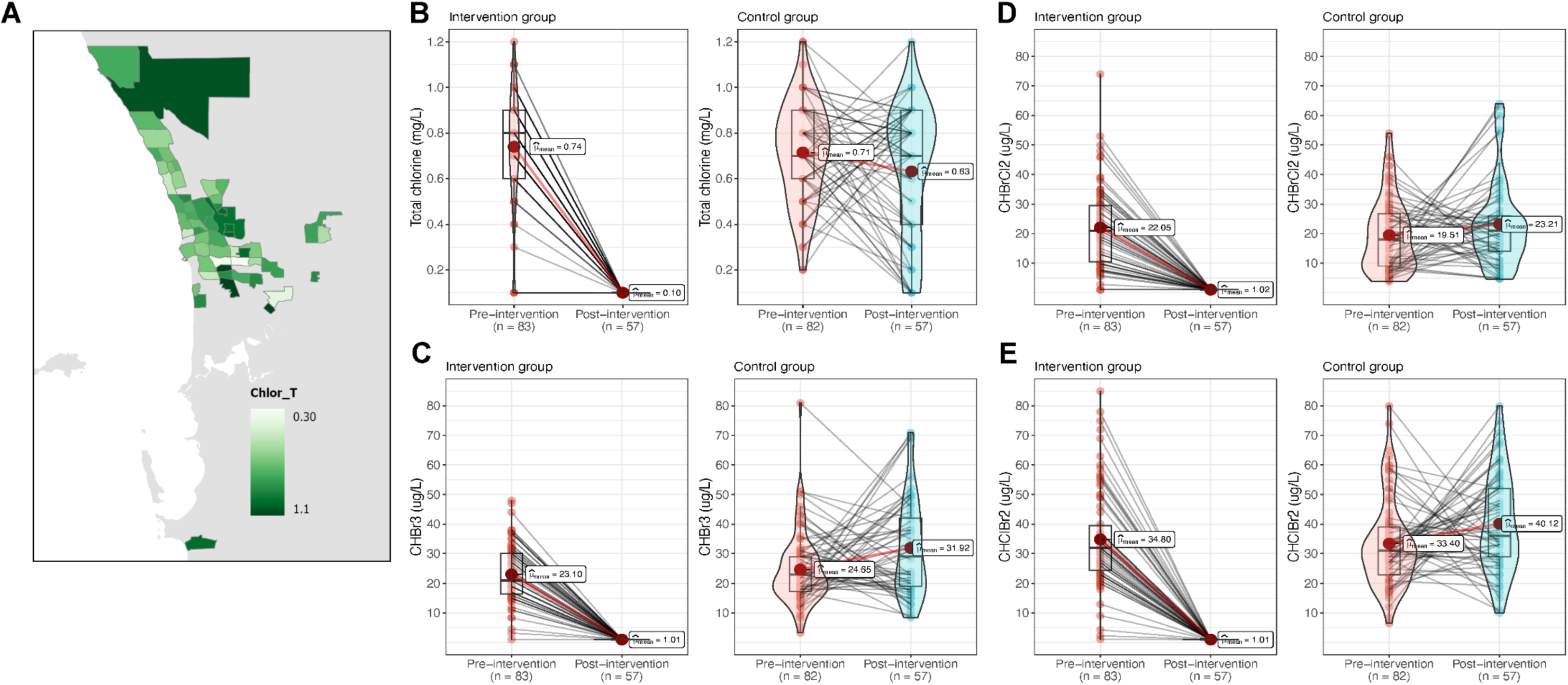
Impact of the intervention on water quality in TUMS participant homes. A geospatial heatmap map displaying mean total chlorine levels (mg/mL), grouped by suburb. Total n=197 (**A**). Boxplot comparison between mean levels of total chlorine (**B**) and disinfection by-products: bromoform (**C**), bromodichloromethane (**D**), and chlorodibromomethane (**E**) between the treatment and control groups. The box indicates the interquartile range (25th to 75th percentile), the horizontal line within the box represents the median, and the whiskers extend to 1.5 times the interquartile range. Outliers are shown as individual points. Total n=140.

The intervention significantly reduced total chlorine levels in the treatment group from a mean of 0.74 mg/L (SD = 0.2447) pre-intervention to 0.10 mg/L (SD = 0.000) post-intervention (paired t-test, p = 2.2^-16^). In contrast, the control group showed no significant change in mean total chlorine levels (0.71 mg/L pre-intervention vs. 0.63 mg/L post-intervention, paired t-test, p = 0.06). The intervention also significantly reduced chlorine disinfection by-products including bromoform, bromodichloromethane, and chlorodibromomethane levels, while the control group showed no significant changes in these disinfection by-products (Figures 1C, 1D, and 1E).

Participant compliance with the intervention, assessed through three monthly surveys, revealed no significant differences between the treatment and control groups. Survey questions encompassed the daily consumption of filtered water (cups per day) by the infant (mean in control group = 3.1, mean in intervention = 2.9, p = 0.06, Welch t-test), usage of study-provided filters outside the home (never, sometimes, always, p = 0.574, Chi-Squared test), and the age at which unboiled tap water was introduced (mean in control group = 10.71, mean in intervention = 10.60, p = 0.781, Welch t-test). These findings suggest consistent adherence to the intervention protocol across both groups (Supplementary Figure 4).

### Microbial community profiling

We analysed 170 stool samples collected at baseline (six months of age) and 130 samples collected at end of intervention (eighteen months of age), totalling 300 unique faecal metagenomes. Batch effect analysis did not identify any significant differences in microbial community composition between sequencing batches (Supplementary Figure 5).

After quality filtering and rarefaction to 7,000,000 read pairs per sample, a total of 3,936,112,872 reads were obtained (mean per sample: 12,990,472 ± 301,687 SD). On average, 47,087 human DNA reads per sample (SD ± 655,372) were removed before analysis. Across all samples, we identified 1,305 bacterial SGBs classified at the species level, belonging to 420 genera, 98 families, 42 orders, 17 classes, and 14 phyla. Additionally, we identified two archaeal SGBs (*Methanobrevibacter smithii* and *Methanobrevibacter smithii_A*) and ten eukaryotic SGBs (various *Candida*, *Clavispora*, *Geotrichum*, *Kluyveromyces*, *Saccharomyces*, *Wickerhamiella*, and *Blastocystis* species). The three dominant phyla across both timepoints and randomisation groups were Actinobacteria, Firmicutes_A, and Bacteroidata and the three dominant species were *Bifobacterium longum*, *Bifobacterum breve*, and *Bifobacterium infantis* (Supplementary Figure 6).

### Effect of chlorinated water on microbiota taxonomic structure and diversity

The taxonomic characterization of participants’ microbiota aligned with previous reports on young infants, with *Bifidobacterium infantis*, *Bifidobacterium longum*, and *Bifidobacterium breve* as the predominant species at six months of age ^29,30^. Interindividual variation in microbiome composition decreased from baseline to end of intervention (Supplementary Figure 7). While both groups at each timepoint displayed similar broad phylum-level patterns (Figure 2A), these changed overall from baseline to end of intervention. Compositional profiles changed significantly from baseline to end of intervention, as captured by beta-diversity metrics and differential abundance testing below.

**Figure 2.**
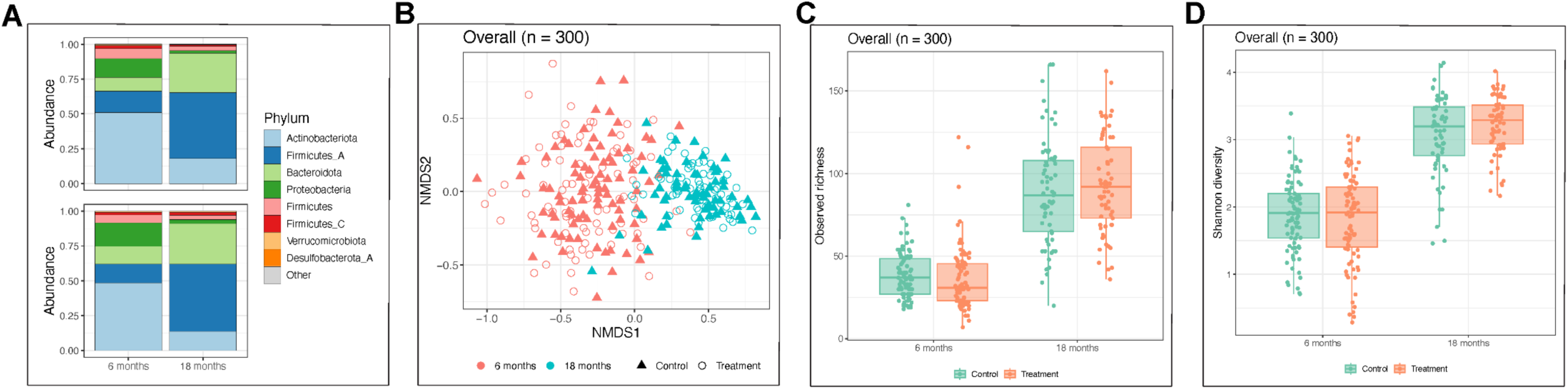
No effect of chlorinated drinking water on the taxonomic structure and diversity of the infant gut microbiome. (**A**) Average relative abundance of bacterial phyla across all samples, stratified by age (six and eighteen months) and treatment group (chlorinated or de-chlorinated water). (**B**) Non-metric multidimensional scaling (NMDS) plot visualizing beta diversity (inter-sample dissimilarity) based on Bray-Curtis distances. Each point represents a sample, coloured by age and shaped by treatment group. (**C**) Boxplots comparing observed richness (number of unique genera) between treatment groups at six and eighteen months of age. (**D**) Boxplots comparing Shannon diversity indices (a measure of both richness and evenness) between treatment groups at six and eighteen months of age. The boxes indicate the interquartile range (25th to 75th percentile), the horizontal line within the box represents the median, and the whiskers extend to 1.5 times the interquartile range. Outliers are shown as individual points.

Beta diversity analysis revealed significant variation associated with age (11.3% of variance explained, PERMANOVA, p = 0.001), but no significant clustering by treatment group (0.6% of variance explained, p = 0.35) in both longitudinal (Figure 2B) and cross-sectional analyses (Supplementary Figure 8). Alpha diversity assessed by richness did not differ significantly between groups before (1.35; 95% CI -3.77-6.47; p=0.6) or after (-4.25; 95% CI: -14.85 - 6.35; p=0.43) treatment (Figure 2C). Similarly, alpha diversity assessed by Shannon Index did not differ significantly between groups before (0.06; 95% CI -0.13 - 0.24; p=0.54) or after (-0.14; 95% CI -0.32 - 0.04; p=0.12) treatment (Figure 2D

### Water chlorination-induced shifts in taxa abundance

Differential abundance analysis using DESeq2 revealed 55 genera across 7 phyla were significantly different in abundance between the intervention (de-chlorinated water) and control (chlorinated water) groups in the eighteen-month (post-intervention) stool samples (adjusted p-value < 0.01) (Figure 3). These differentially abundant genera were primarily members of the Firmicutes phylum. However, due to high inter-individual variation, most of these taxa were present at low frequencies in both groups, although they were more frequently detected in the intervention group (Supplementary Figure 9). 13 genera across 5 phyla were initially different between the intervention and control groups at baseline (Supplementary Figure 10A). A comprehensive list of all differentially abundant SGBs, along with their statistical significance, is provided in Supplementary Table 1.

**Figure 3.**
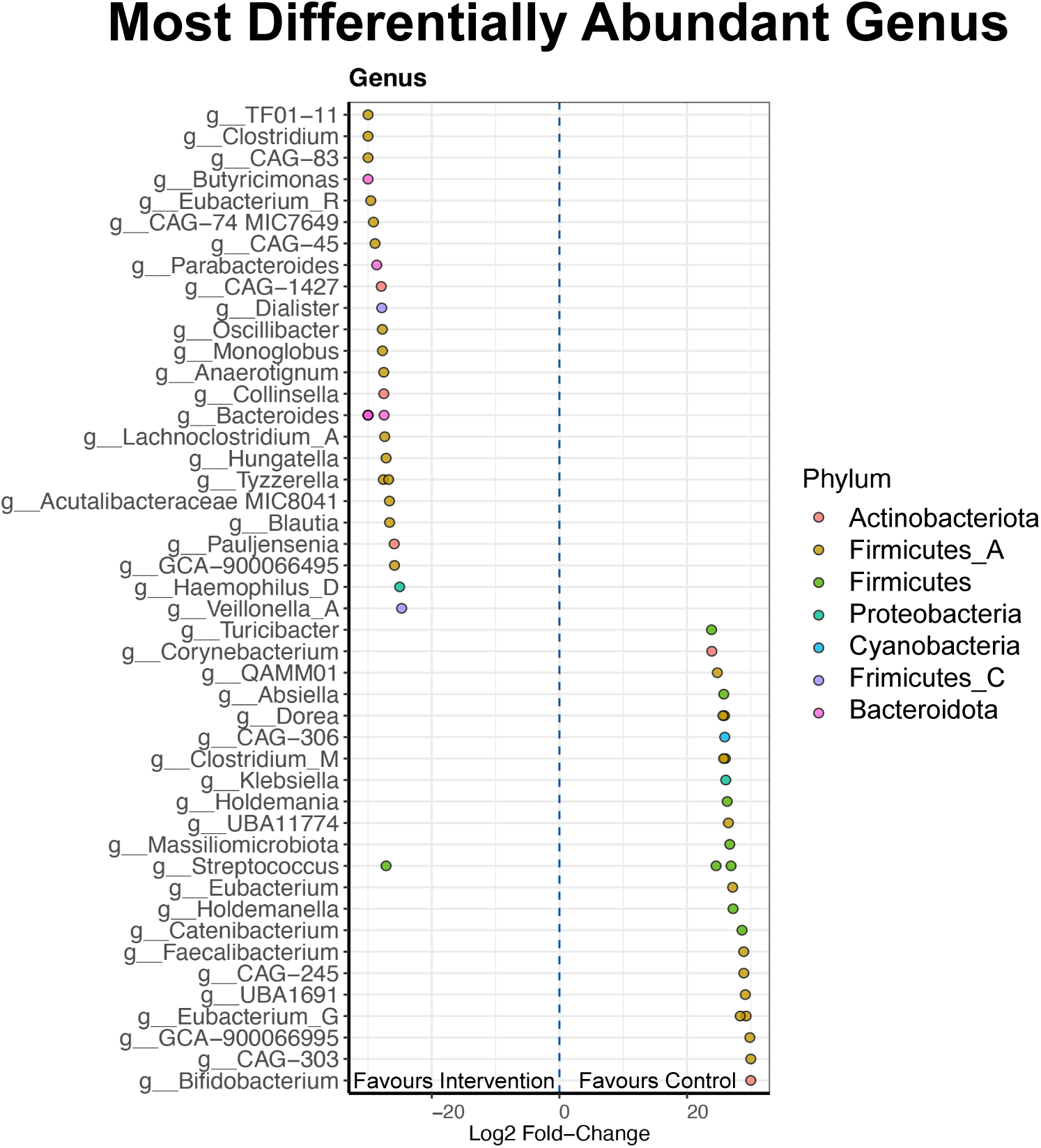
Differential abundance of bacterial genera in infant gut microbiota between treatment groups. The dot plot displays the log2 fold change in abundance for each bacterial genus identified as significantly different (adjusted p-value < 0.01) between the de-chlorinated water (intervention) and chlorinated water (control) groups at eighteen months of age. Each dot represents a genus, coloured by phylum. A positive log2 fold change indicates higher abundance in the intervention group, while a negative value indicates higher abundance in the control group. Data are plotted at the genus level for ease of visualisation.

### Higher antibiotic resistance gene abundance in children receiving chlorinated water

To assess the impact of water chlorination on gut microbiome function, we analysed the abundance of metabolic pathways annotated in the MetaCyc database using DESeq2 ^28^. In the eighteen-month (post-intervention) samples, we identified 901 metabolic pathways belonging to 51 major groups. Two of these groups were differentially abundant between the intervention and control, with chlorinated water exposure most significantly associated with higher abundance of the ’Antibiotic Resistance’ major group, as well as the sundry group ‘Other’ (adjusted p < 0.05) (Figure 4A, Table 2). At the pathway level, 29 metabolic pathways showed higher relative abundance in the chlorinated water group (adjusted p < 0.05), including polymyxin resistance which is a member of the ’Antibiotic Resistance’ major group (Figure 4B, Table 2), which was not significantly different at baseline (Supplementary Figure 10B, Supplementary Table 2). There were no significantly differentially abundant MetaCyc major groups between the intervention and control group at baseline. To determine which species contributed to the higher abundance of antibiotic resistance pathways, we examined the species abundance in individual microbiomes that were annotated to the MetaCyc ’Antibiotic Resistance’ functional pathway. This analysis revealed that antibiotic resistance genes were distributed across multiple species, including *Escherichia*, *Klebsiella*, *Enterobacter*, *Clostridia*, and *Citrobacter*, among others (Supplementary Figure 11).

**Figure 4.**
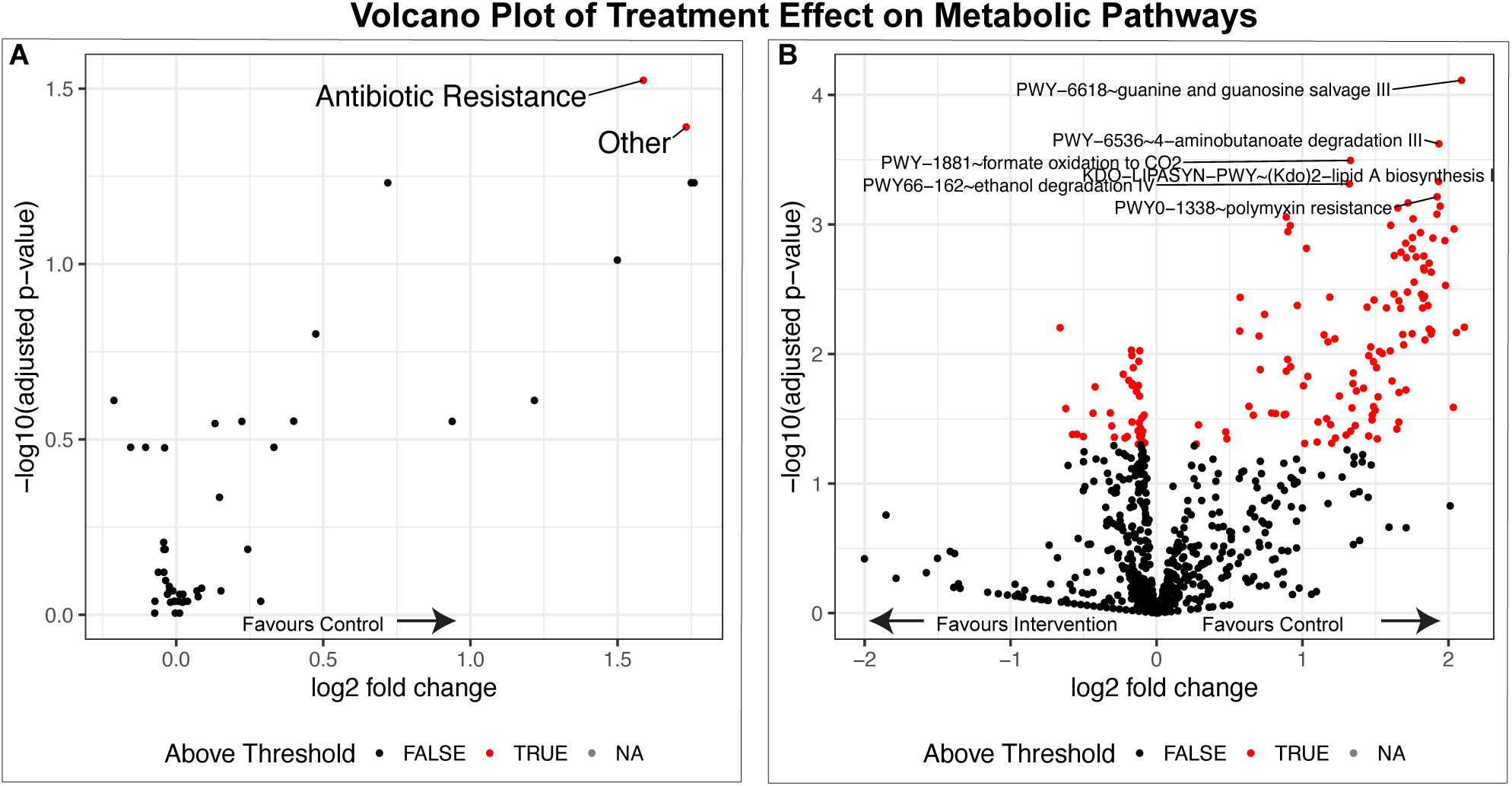
Impact of water chlorination on infant gut microbiome metabolic pathways (post-intervention). (**A**) Volcano plot displaying the differential abundance of MetaCyc metabolic groups between the de-chlorinated water (intervention) and chlorinated water (control) groups. Each point represents a metabolic group, with the x-axis showing log2 fold change and the y-axis showing -log10 adjusted p-value. Red points indicate groups with significantly higher abundance in the control group (adjusted p < 0.05). (**B**) Volcano plot displaying the differential abundance of individual MetaCyc metabolic pathways between the intervention and control groups, following the same format as panel **A**.

**Table 2.** Significantly differentially abundant MetaCyc groups (p<0.05) and pathways (adjusted p<0.05) between the intervention and control groups at end of intervention (rounded to four decimal places)

| MetaCyc major group | Log2FoldChange | p-value | Adjusted p-value |
| --- | --- | --- | --- |
| Antibiotic Resistance | 1.5876 | 0.0006 | 0.0300 |
| Other | 1.7332 | 0.0017 | 0.0407 |
| Methylglyoxal Detoxification | 1.7602 | 0.0058 | 0.0587 |
| TCA cycle | 1.7502 | 0.0060 | 0.0587 |
| Respiration | 0.7191 | 0.0052 | 0.0587 |
| Entner-Duodoroff Pathways | 1.4993 | 0.0119 | 0.0975 |
| Fatty Acid and Lipid Degradation | 0.47441 | 0.0226 | 0.1583 |
| Metabolic Regulator Biosynthesis | -0.2116 | 0.0430 | 0.2448 |
| O-Antigen Biosynthesis | 1.2175 | 0.0450 | 0.2448 |
| **MetaCyc pathway** |  |  |  |
| PWY-6618~guanine and guanosine salvage III | 0.5285 | 0.0001 | 0.0480 |
| PWY-6536~4-aminobutanoate degradation III | 0.5265 | 0.0002 | 0.0480 |
| PWY-1881~formate oxidation to CO2 | 0.3693 | 0.0003 | 0.0480 |
| KDO-LIPASYN-PWY~(Kdo)2-lipid A biosynthesis I | 0.5522 | 0.0005 | 0.0480 |
| PWY66-162~ethanol degradation IV | 0.3786 | 0.0005 | 0.0480 |
| PWY0-1338~polymyxin resistance | 0.5610 | 0.0006 | 0.0480 |
| PWY-6537~4-aminobutanoate degradation II | 0.5072 | 0.0007 | 0.0480 |
| PWY0-1471~uracil degradation III | 0.5748 | 0.0007 | 0.0480 |
| GLUCARGALACTSUPER-PWY~superpathway of D-glucarate and D-galactarate degradation | 0.4901 | 0.0007 | 0.0480 |
| PWY-6896~thiamine salvage I | 0.5748 | 0.0008 | 0.0480 |
| PWY-46~putrescine biosynthesis III | 0.2671 | 0.0009 | 0.0480 |
| CHOLINE-BETAINE-ANA-PWY~choline degradation I | 0.5296 | 0.0009 | 0.0480 |
| BETSYN-PWY~glycine betaine biosynthesis I (Gram-negative bacteria) | 0.5296 | 0.0009 | 0.0480 |
| PWY66-394~aspirin triggered resolvin E biosynthesis | 0.4882 | 0.0010 | 0.0480 |
| P283-PWY~hydrogen oxidation I (aerobic) | 0.2789 | 0.0010 | 0.0480 |
| SO4ASSIM-PWY~assimilatory sulfate reduction I | 0.6237 | 0.0011 | 0.0480 |
| PWY-6050~dimethyl sulfoxide degradation | 0.2765 | 0.0011 | 0.0480 |
| TCA-GLYOX-BYPASS~superpathway of glyoxylate bypass and TCA | 0.5565 | 0.0012 | 0.0480 |
| FERMENTATION-PWY~mixed acid fermentation | 0.5438 | 0.0013 | 0.0480 |
| PWY-6535~4-aminobutanoate degradation I | 0.5876 | 0.0013 | 0.0480 |
| DETOX1-PWY-1~reactive oxygen species degradation | 0.6156 | 0.0013 | 0.0480 |
| PWY-7417~phospholipid remodeling (phosphatidate, yeast) | 0.5341 | 0.0014 | 0.0480 |
| PWY0-1241~ADP-L-glycero-β-D-manno-heptose biosynthesis | 0.3240 | 0.0015 | 0.0480 |
| PWY0-981~taurine degradation IV | 0.5531 | 0.0015 | 0.0480 |
| PUTDEG-PWY~putrescine degradation I | 0.5320 | 0.0016 | 0.0480 |
| PWY-5137~fatty acid β-oxidation III (unsaturated, odd number) | 0.5200 | 0.0017 | 0.0480 |
| TCA~TCA cycle I (prokaryotic) | 0.5852 | 0.0018 | 0.0480 |
| PWY-5755~4-hydroxybenzoate biosynthesis II (bacteria) | 0.5693 | 0.0018 | 0.0480 |
| PWY-7856~heme a biosynthesis | 0.5484 | 0.0018 | 0.0480 |

## DISCUSSION

In this double-blinded randomised controlled trial conducted in a high-income setting, infants exposed to chlorinated drinking water exhibited no differences in the overall richness and diversity of the microbiota, although subtle shifts in taxonomic abundance were observed. Several species known to be part of a healthy gut microbiome, including *Bacteroides, Clostridium,* and *Blautia* showed reductions in taxonomic abundance. While some beneficial bacteria like *Faecalibacterium prauznitzii* and *Bifidobacterium* increased in abundance, others including *Streptococcus, Clostridia* and *Klebsiella pneumoniae* also increased, which could be opportunistic pathogens under certain circumstances. It is important to note that carriage of differentially abundant taxa was generally low across the cohort against a background of strikingly individualized microbiota compositions in this age group. On balance, our data suggest chlorination-induced shifts in the microbiota vary substantially from infant to infant, possibly due to a range of factors including genetics, diet and environmental exposures.

Consistent with the recent study by Nadimpalli et al. ^13^, we found that exposure to chlorinated drinking water for a twelve-month period was associated with a higher carriage of antibiotic resistance genes in stool samples. These ARGs, which encompass genes encoding efflux pumps and antibiotic-degrading enzymes, were detected across diverse bacterial species, including *Escherichia*, *Klebsiella*, *Enterobacter*, *Clostridia*, and *Citrobacter*. Taken together, our study and the Bangladeshi trial suggests chlorinated water may selectively promote the growth of bacteria harbouring ARG genes ^13^. It is noteworthy that the age group in the previous study where chlorination effects were observed (15-30 months) overlaps with the age range of our study participants. This raises the possibility of a developmental window during which the gut microbiome may be particularly susceptible to environmental exposures such as chlorinated water. However, we acknowledge that the observed changes in microbial composition may reflect a relative enrichment of more chlorine-tolerant species, rather than direct promotion of their growth. Importantly, the chlorination levels in our study (0.3–1.1 mg/L) are substantially lower than those used in the Bangladeshi trial, yet we observed comparable shifts in microbial composition and ARG carriage. This suggests that even low-level chlorination, typical of public water systems in high-income countries, may influence the developing gut microbiome and resistome. Further investigation is required to ascertain the potential for the development or spread of antibiotic resistance in this population.

These findings have important public health implications. Antimicrobial resistance (AMR) is a growing global health threat, identified by the World Health Organization as one of the top global public health priorities ^2^. While efforts to combat AMR have largely focused on reducing antibiotic use and promoting vaccination, our study suggests that environmental exposures, such as residual disinfectants in drinking water, may also play a role in shaping the resistome in early life. Children receive frequent antibiotics in the first five years, which may interact with a chlorinated-water driven ARG gene expression in undesirable ways. Further understanding these interactions is now cruicial for developing comprehensive mitigation strategies.

Our study has limitations, including a relatively small sample size and the inability to draw definitive conclusions about infant health outcomes. Antibiotic use, recorded in approximately one-third of participants equally across both groups, could not be controlled for in a per-protocol analysis due to power limitations. While compliance and water consumption practises were monitored, the potential for unmeasured confounders is acknowledged. We acknowledge that infant feeding method is an important determinant of gut microbiota composition. In our study, the distribution of feeding categories (predominantly breastfed, formula-fed, and mixed fed) was balanced between the treatment and control groups (Table 1). Infant feeding method was recorded at recruitment at six months of age, meaning we were unable to address the role of maternal drinking water exposure during the breastfeeding period. Additionally, our findings cannot be solely attributed to chlorine, as the study filters also removed nitrates, heavy metals, and disinfection by-products, however, among these components chlorine and its by-products remain the most plausible candidates given their well-established antimicrobial properties.

Although all measured chlorine levels in our study fell below the 5 mg/L limit set by the Australian Drinking Water Guidelines, the range observed (0.3–1.1 mg/L) is typical of residual disinfectant concentrations used in public water systems internationally. Our findings therefore reflect real-world exposures encountered by large populations. However, we acknowledge that even within this range, local variations in water chemistry and treatment practices may influence microbiome outcomes, and caution is warranted in extrapolating these findings beyond comparable settings.

To our knowledge, this is the first double-blinded RCT to examine the impact of chlorinated drinking water on the infant gut microbiome and resistome in a high-income country setting. While similar studies have been conducted in Low- and Middle-Income Countries, the water treatment context, baseline microbiota, and environmental exposures differ substantially, making our findings uniquely relevant to urban populations in developed settings.

## CONCLUSION

Our study extends previous findings that suggest chlorinated drinking water may drive adaptations in the gut microbiota influencing antibiotic resistance gene carriage. Further work is now needed to understand how this might influence health outcomes, particularly among children exposed to frequent antibiotics in early childhood.

## Data Availability

Raw data supporting the conclusions of the article will be make available by the authors upon request.

## DECLARATIONS

### Ethics Approval and Consent to Participate

The study protocol was approved by the Ramsay Health Care (RHC) WA | SA Human Research Ethics Committee (EC00266; HREC reference number 1911). Mothers provided written informed consent on behalf of their children. All procedures were conducted in accordance with the Declaration of Helsinki.

### Consent for Publication

This study does not contain any individual person’s data, only aggregate data are presented.

### Availability of Data

Source data, de-identified participant data, study protocol, analysis code, and other materials supporting the conclusions of this article are available under CC-By Attribution 4.0 International license on the Open Science Framework landing page for this project https://osf.io/krjbe/?view_only=bf3fc85240ed4e6a9324838132837a1a.

The direct link to the Mendeley Data repository: https://data.mendeley.com/datasets/8f59b8sxwv/2

### Authors Contributions

Conceptualization (DM, DS, MC, DP, CC, VV, SP). Data curation (KP). Formal analysis (KP). Writing – original draft (KP, DM). Writing – review & editing (all authors).

## Acknowledgements

The authors gratefully acknowledge the invaluable contributions of the ORIGINS Project families for their ongoing support of The Water Quality and Microbiome Study (TUMS). We extend our sincere thanks to the following teams and individuals who have made The ORIGINS Project possible: The ORIGINS Project team; Joondalup Health Campus (JHC); members of ORIGINS Project Community Reference and Participant Reference Groups; Research Interest Groups and the ORIGINS Project Scientific Committee; The Kids Research Institute Australia; City of Wanneroo; City of Joondalup; and Fiona Stanley.

We are also grateful for the contributions of Richard Theobald (WA Department of Health) for advice on water quality testing and study design considerations; The Kids Research Institute Australia Biostatistics team (Ali Hollingsworth, Zac Dempsey) for their assistance with randomisation, blinding and compliance monitoring; Nikki Schultz for her analysis of the compliance data, the Water Corporation WA (Cameron Gordon, Steve Christie, Gabrielle O’Dwyer) for their interest, advice, and engagement with the study. We acknowledge the support of our research partners, Microba Life Sciences, ChemCentre and Pure Water Systems for their additional in-kind support of the project. We would also like to thank Jen Rozier from The Kids Research Institute Australia for the geospatial mapping analysis, and Cynthia Joll from Curtin University for her expertise and advice on water quality guidelines in Australia. The initial draft of this manuscript was refined for clarity with the assistance of Gemini Advanced, Google’s large language model.

## Notes

### Competing Interest Statement

The authors have declared no competing interest.

### Clinical Trial

ACTRN12619000458134

### Funding Statement

This work was supported by Telethon Kids Institute Blue Sky award 2018. Individual authors are supported by: DM work was supported by the Western Australian Future Health Research and Innovation Fund, which is an initiative of the WA State Government. VV work was supported by the funding endowed by the Family Larsson-Rosenquist Foundation. DJP was supported by the NHMRC Medical Research Futures Fund (MRFF) Career Development Fellowship ID1144544 (2018-2021) and the Telethon Kids Institute Ascend Fellowship (2022-2026).

### Summary of Updates

The revision enhances emphasis on the main novel findings from the study, that chlorinated drinking water, while having minor effects on composition, significantly upregulated ARG gene pathways in children. The results, figures and tables remain unaltered from version 1. Only the framing of the introduction and discussion have changed.

