## Supplemental Figures and Tables for "Chlorinated Drinking Water Exposure Enriches Antimicrobial Resistance Pathways in the Infant Gut Microbiome: A Randomized Trial"

**Table of contents:**

1. Supplementary Figure 1 - Graphical overview of trial design

2. Supplementary Figure 2 - CONSORT flow diagram of participant recruitment, enrolment, allocation, follow-up, and analysis

3. Supplementary Figure 3 - A geospatial map displaying disinfection by-product levels, grouped by suburb, measured prior to intervention

4. Supplementary Figure 4 - Analysis of compliance surveys administered quarterly suggests similar adherence between groups

5. Supplementary Figure 5 - Non-metric multidimensional scaling analysis revealed no evidence of batch effect between sequencing runs

6. Supplementary Figure 6 - Taxa bar plots showing the dominant phyla and species across both timepoints and randomisation groups

7. Supplementary Figure 7 - Taxa bar plot showing higher rates of inter-individual variation at baseline compared to end of intervention

8. Supplementary Figure 8 - NMDS analysis of beta diversity

9. Supplementary Figure 9 - Box plots of log-transformed counts of top-ranked differentially abundant species

10. Supplementary Figure 10 - Impact of water chlorination on infant gut microbiome metabolic pathways and differential abundance of bacterial genera at baseline

11. Supplementary Figure 11 - Heatmap of sequencing read counts by species

13. Supplementary Table 1

14. Supplementary Methods

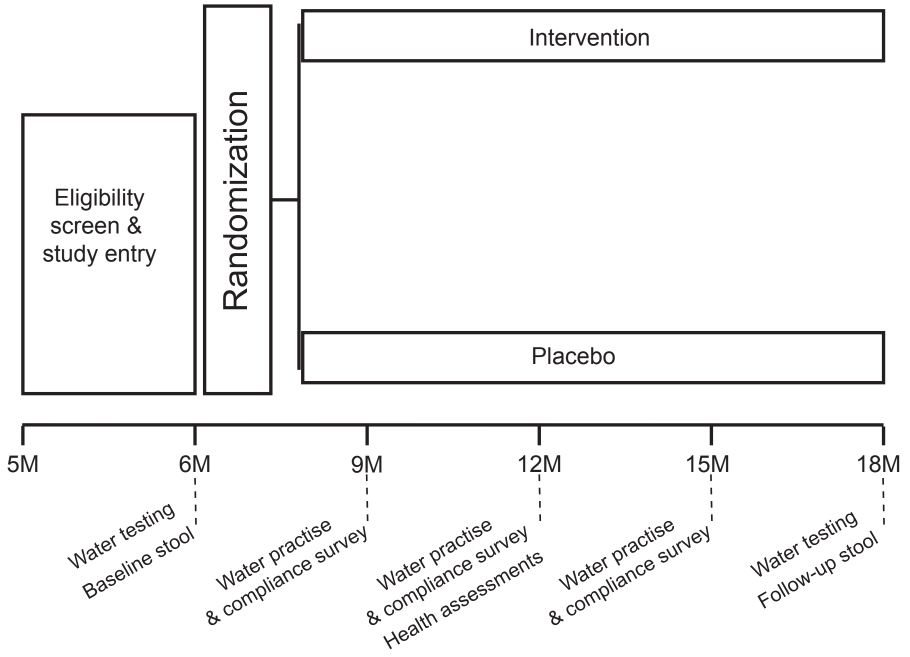


**Supplementary Figure 1 – Graphical overview of trial design.** A timeline of data collection events is shown as a function of infant age in months.

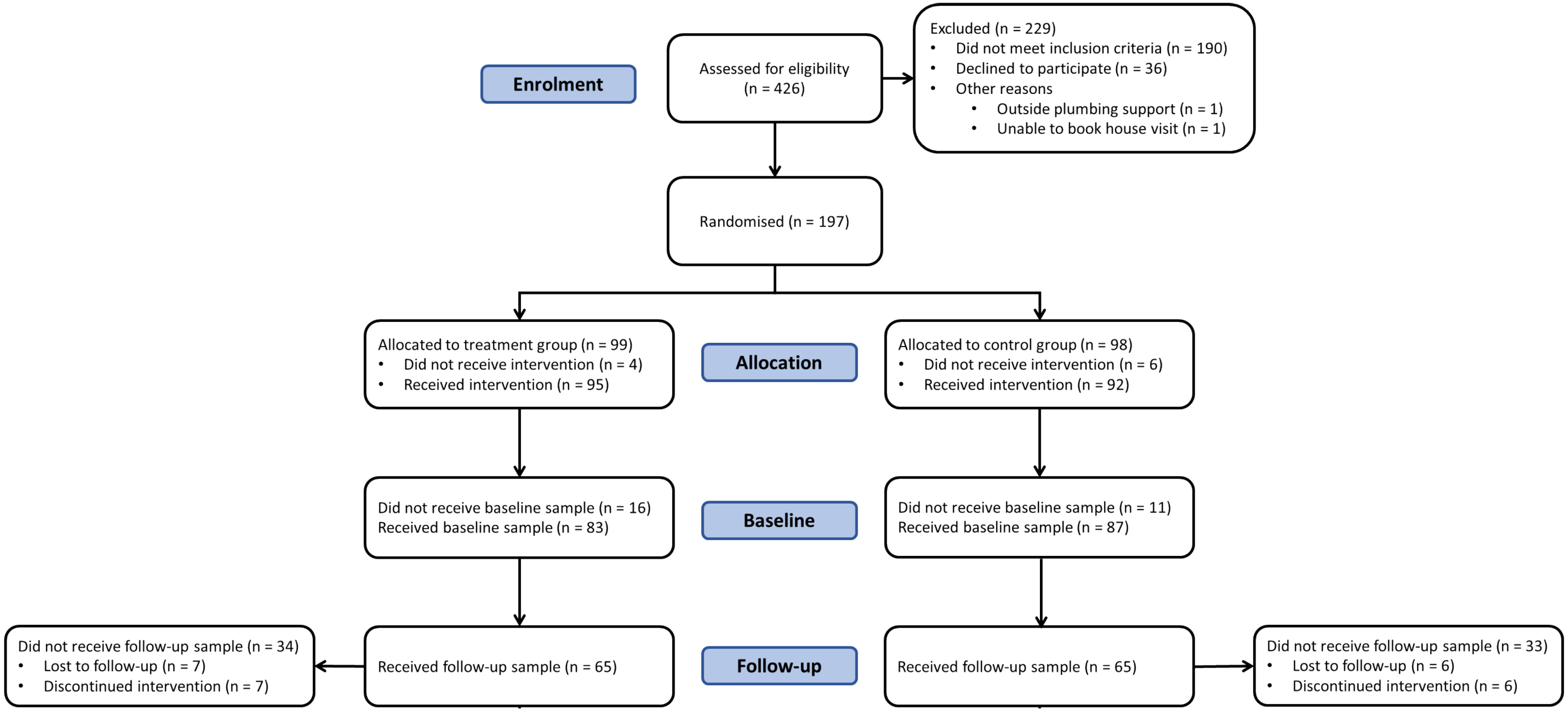


**Supplementary Figure 2 - CONSORT flow diagram of participant recruitment, enrolment, allocation, follow-up, and analysis.** This diagram illustrates the flow of participants through the different stages of the TUMS randomised controlled trial. It shows the number of infants assessed for eligibility, reasons for exclusion, randomisation into the intervention and control groups, loss to follow-up, and the final number of participants included in the analysis.

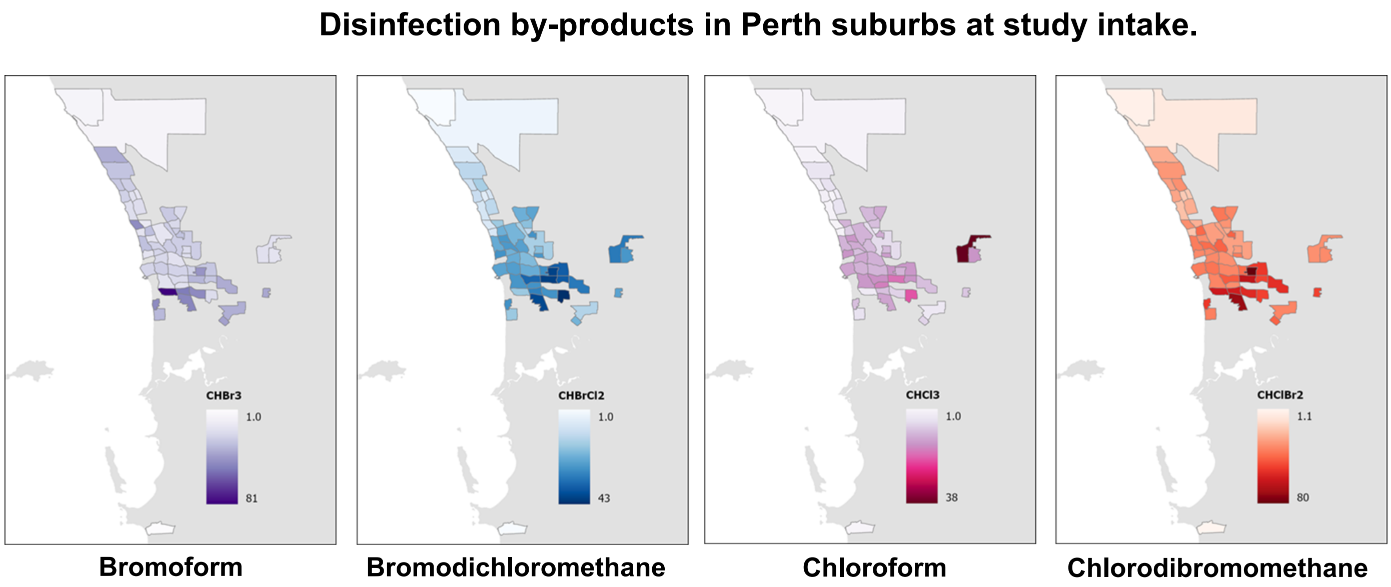


**Supplementary Figure 3 – A geospatial map displaying disinfection by-product levels, grouped by suburb, measured prior to intervention.** Visualisation represents DBPs expressed as ug/mL averaged over participant homes and represented at the suburb level.

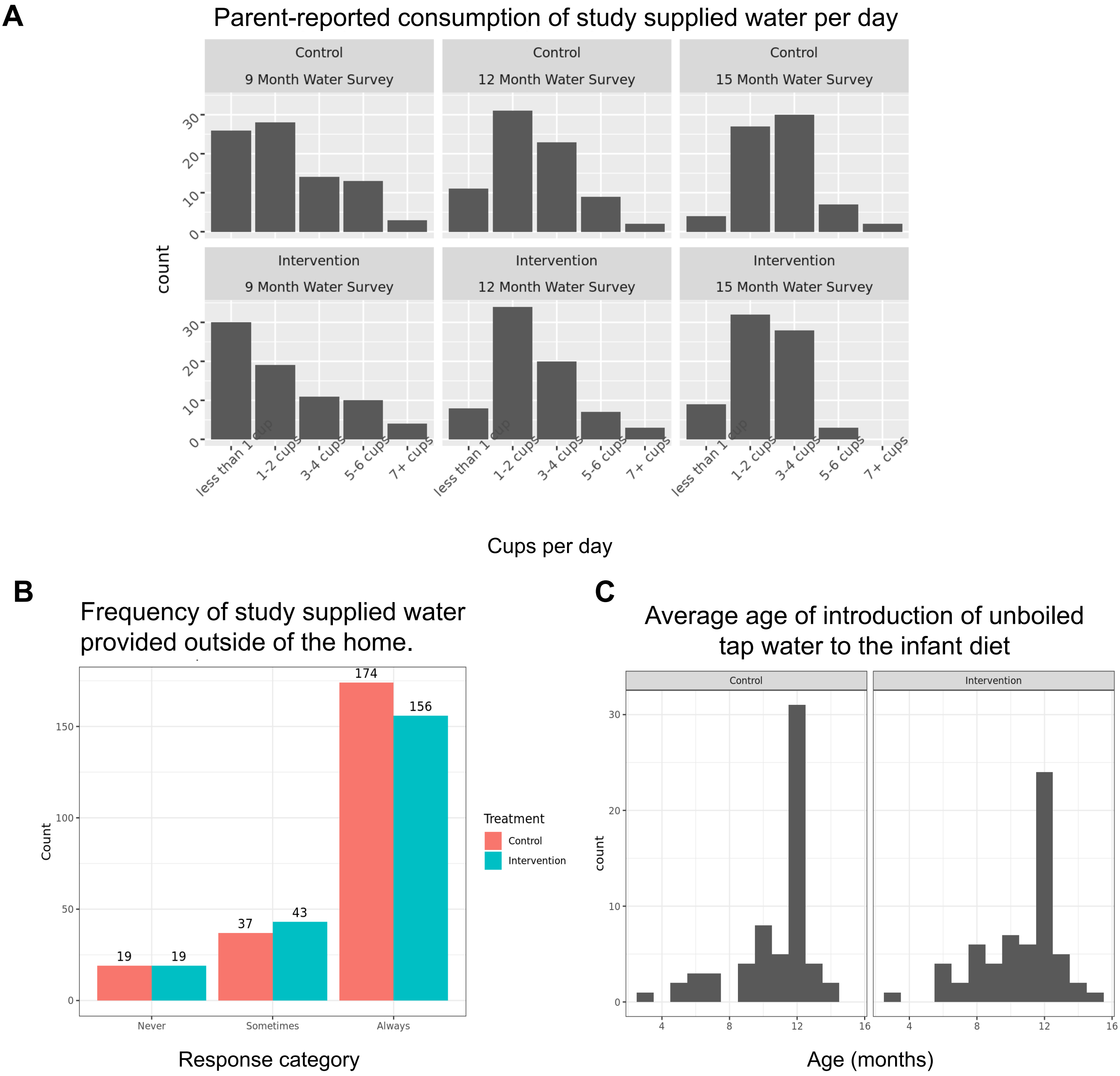


**Supplementary Figure 4 – Analysis of compliance surveys administered quarterly suggests similar adherence between groups. (A)** Frequency count of parent-reported consumption of study supplied water per day, measured by quarterly questionnaires exhibited a similar distribution between groups. **(B)** Frequency count of consumption of study supplied water when outside the home, using supplied water bottles was similar across categories. **(C)** Distribution of parent reported age of introduction of tap water into the infant diet averaged over surveys was similar between groups.

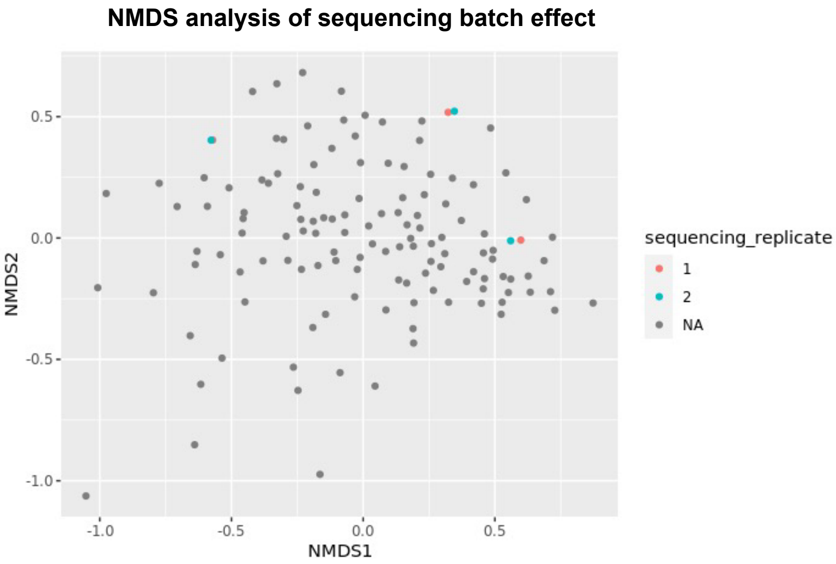


**Supplementary Figure 5 – Non-metric multidimensional scaling analysis revealed no evidence of batch effect between sequencing runs.** Stool samples were sequenced in two batches. The first batch included all 170 baseline samples and 35 end of intervention samples. The remaining end of intervention samples, along with three end of intervention samples re-sequenced from the first batch, were included in the second batch. Non-metric multidimensional scaling (NMDS) ordination demonstrates no significant difference in gut microbial community composition between sequencing batches. Each point represents an individual sample. Red symbols denote three randomly selected samples sequenced in batch 1, while blue symbols represent the same three samples re-sequenced in batch 2. The proximity of red and blue symbols for each sample indicates minimal variation introduced by sequencing batch.

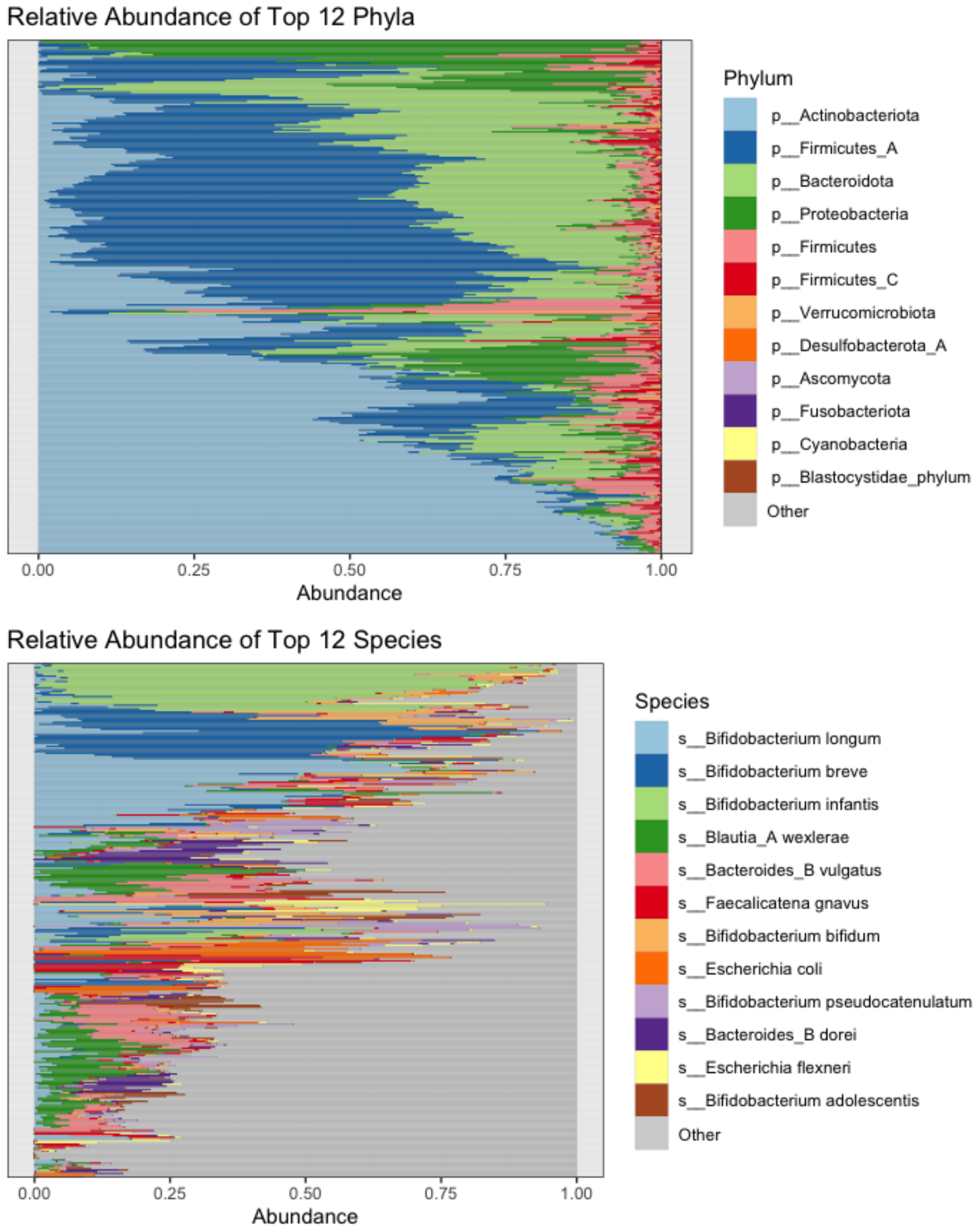


**Supplementary Figure 6 - Taxa bar plots showing the dominant phyla ans species across both timepoints and randomisation groups.** Taxonomic bar plot illustrating the top twelve most abundant phyla (top) and species (bottom), ranked by Bray-Curtis dissimilarity. Each bar represents an individual participant, coloured according to the relative abundance of each genus (x-axis).

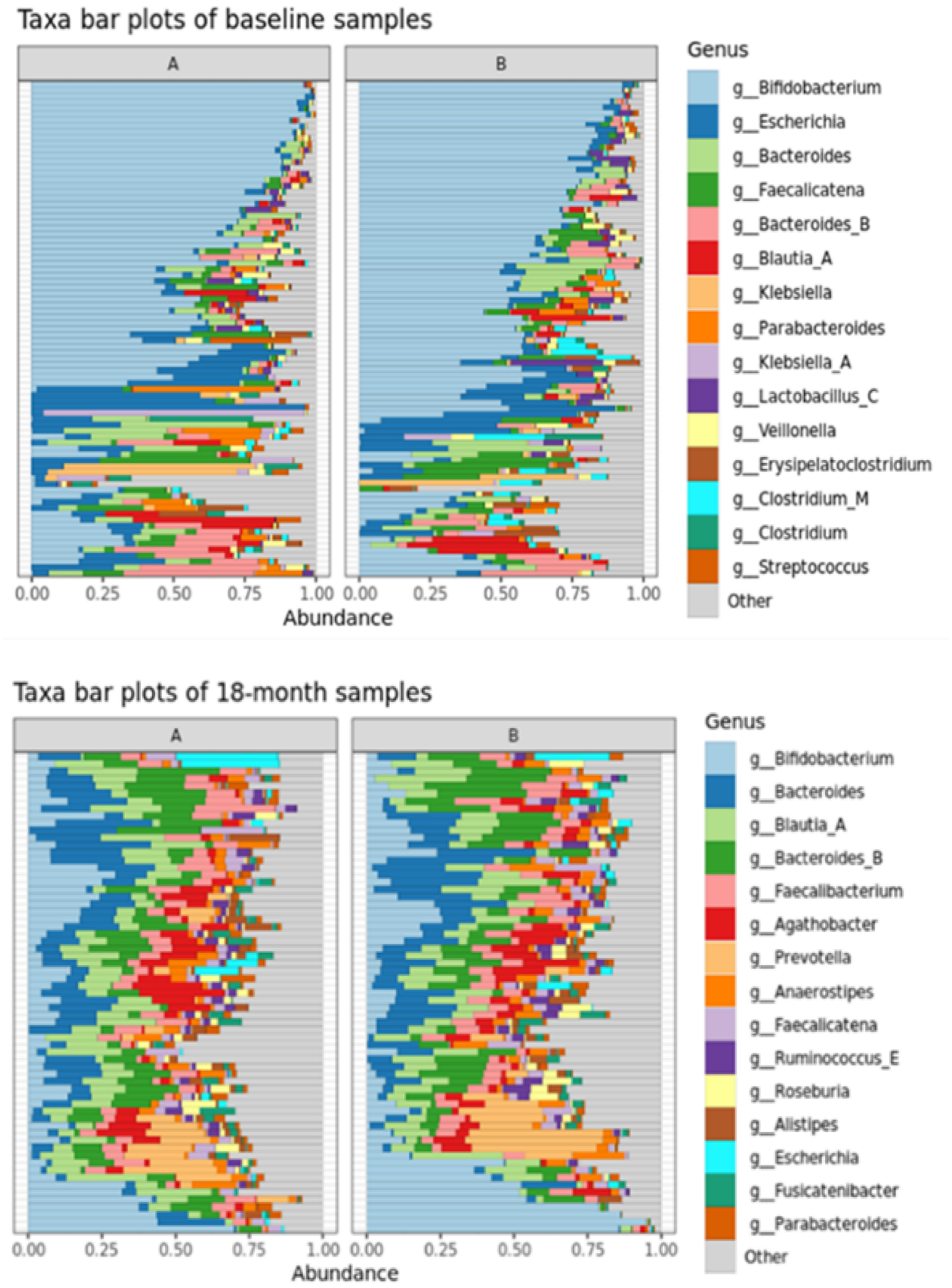


**Supplementary Figure 7 - Taxa bar plot showing higher rates of inter-individual variation at baseline compared to end of intervention.** Taxonomic bar plot illustrating the top twelve most abundant genera at baseline (top) and at end of intervention (bottom), ranked by Bray-Curtis dissimilarity within each time point. Each bar represents an individual participant, coloured according to the relative abundance of each genus (x-axis).

NMDS analysis of treatment main effect

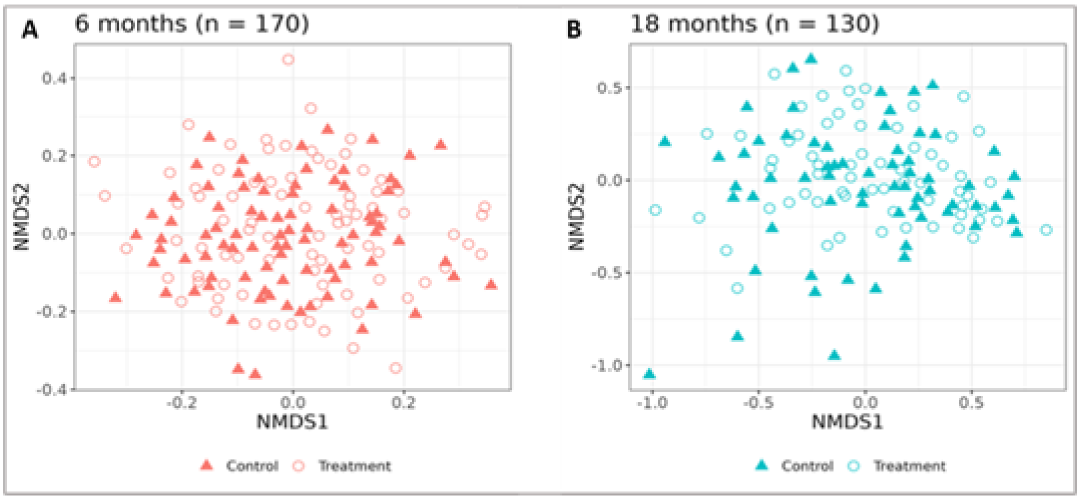


**Supplementary Figure 8 – NMDS analysis of Beta Diversity.** A two-dimensional representation of the pairwise genomic distances between each sample, as identified by NMDS analysis. Pairwise distances when stratified by child age: six months (**A**), and eighteen months (**B**).

**Top Four Most Differentially Abundant Species**

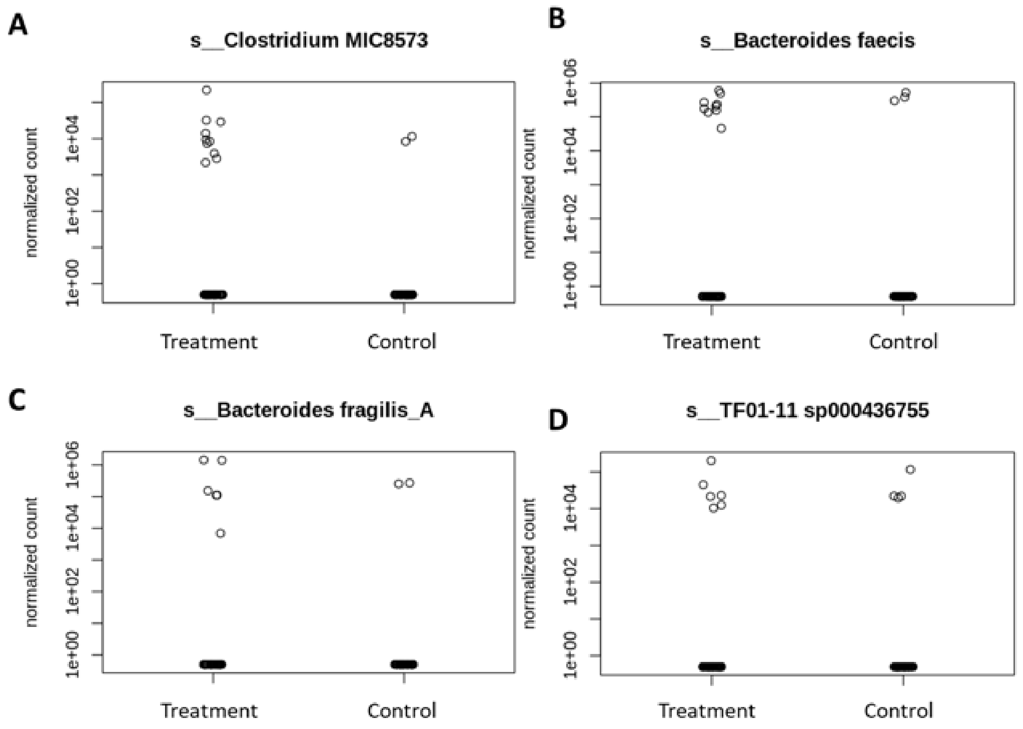


**Supplementary Figure 9 – Box plots of log-transformed counts of top-ranked differentially abundant species.** Box plots display the log-transformed abundance of the top four Amplicon Sequence Variants (ASVs) exhibiting significant differences between the intervention and control groups: (**A**) *Clostridium MIC8573*, (**B**) *Bacteroides faecis*, (**C**) *Bacteroides fragilis_A*, and (**D**) *TF01-11 sp000436755*.

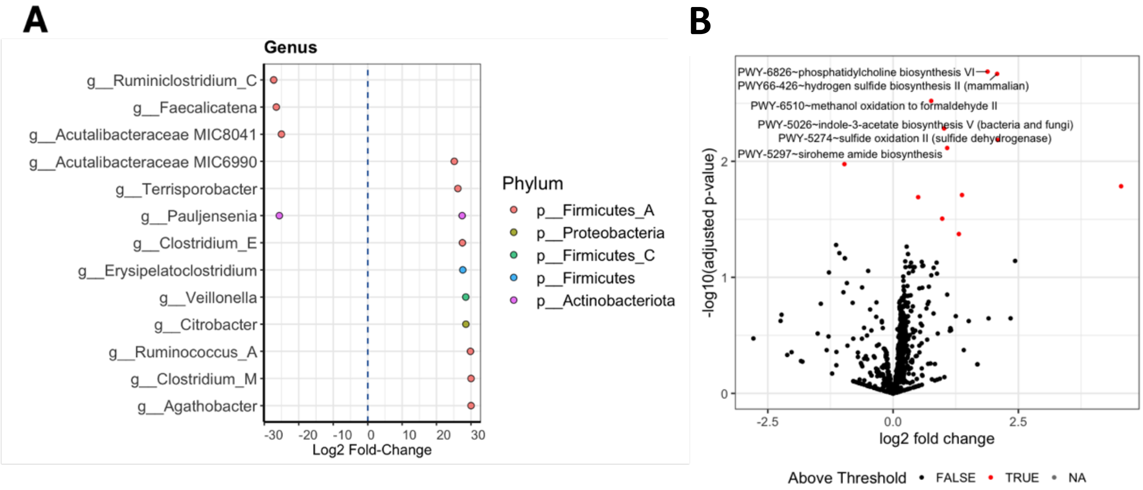


**Supplementary Figure 10. Impact of water chlorination on differential abundance of bacterial genera and infant gut microbiome metabolic pathways at baseline.** (**A**) Dot plot displaying the log2 fold change in abundance for each bacterial genus identified as significantly different (adjusted p-value < 0.01) between the de-chlorinated water (intervention) and chlorinated water (control) groups at baseline. Each dot represents a genus, coloured by phylum. A positive log2 fold change indicates higher abundance in the intervention group, while a negative value indicates higher abundance in the control group. Data are plotted at the genus level for ease of visualisation. (**B**) Volcano plot displaying the differential abundance of individual MetaCyc metabolic pathways between the intervention and control groups.

### Log2 Sequencing reads for species annotated to the Antibiotic Resistance Metacyc pathway

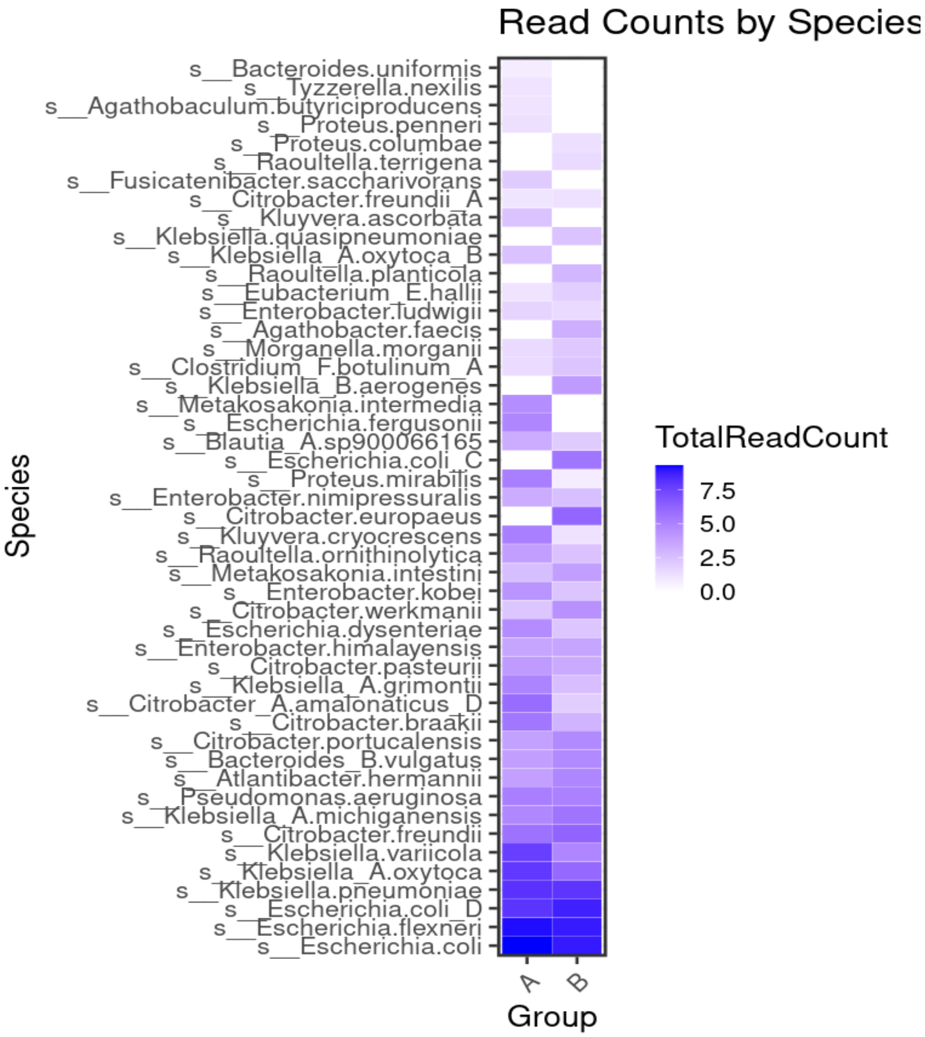


**Supplementary Figure 11 – Heatmap of Sequencing Read Counts by Species.** Heatmap depicting the abundance of bacterial species harbouring genes associated with antibiotic resistance (as defined by the MetaCyc database) within the gut microbiome of each infant. Rows represent individual bacterial species, while columns represent treatment groups. Cell colour intensity reflects the relative abundance of each species within the respective group, highlighting potential differences in antibiotic resistance profiles among infants receiving different treatments. A = intervention, B = control.

**Supplementary Table 1: Differentially abundant species between the intervention (de-chlorinated water) and control (chlorinated water) groups at end of intervention.**

| Species | log2FoldChange | p-value | Adjusted p-value | Effect of chlorinated water |
|---|---|---|---|---|
| *Clostridium MIC8573* | -30.00 | 1.34e-29 | 4.46e-27 | Reduces abundance |
| *Bacteroides faecis* | -30.00 | 4.63e-28 | 7.69e-26 | Reduces abundance |
| *Bacteroides fragilis_A* | -30.00 | 3.87e-25 | 1.43e-23 | Reduces abundance |
| *TF01-11 sp000436755* | -30.00 | 3.87e-25 | 1.43e-23 | Reduces abundance |
| *Bacteroides clarus* | -30.00 | 3.87e-25 | 1.43e-23 | Reduces abundance |
| *CAG-83 sp000435555* | -30.00 | 3.87e-25 | 1.43e-23 | Reduces abundance |
| *Butyricimonas synergistica_A* | -30.00 | 3.87e-25 | 1.43e-23 | Reduces abundance |
| *Eubacterium_R sp003526845* | -29.57 | 1.81e-24 | 5.45e-23 | Reduces abundance |
| *CAG-74 MIC7649* | -29.15 | 8.10e-24 | 1.94e-22 | Reduces abundance |
| *CAG-45 sp000438375* | -28.90 | 1.91e-23 | 3.87e-22 | Reduces abundance |
| *Parabacteroides goldsteinii* | -28.63 | 4.80e-23 | 8.43e-22 | Reduces abundance |
| *CAG-1427 sp000435675* | -27.93 | 5.31e-22 | 8.39e-21 | Reduces abundance |
| *Dialister MIC7247* | -27.85 | 6.92e-22 | 1.04e-20 | Reduces abundance |
| *Oscillibacter MIC7169* | -27.75 | 9.55e-22 | 1.38e-20 | Reduces abundance |
| *Monoglobus pectinilyticus* | -27.73 | 1.02e-21 | 1.41e-20 | Reduces abundance |
| *Tyzzerella MIC7714* | -27.59 | 1.63e-21 | 2.16e-20 | Reduces abundance |
| *Anaerotignum sp001304995* | -27.53 | 1.99e-21 | 2.48e-20 | Reduces abundance |
| *Collinsella sp003487125* | -27.53 | 2.02e-21 | 2.48e-20 | Reduces abundance |
| *Bacteroides nordii* | -27.48 | 2.37e-21 | 2.80e-20 | Reduces abundance |
| *Lachnoclostridium_A edouardi* | -27.38 | 3.33e-21 | 3.81e-20 | Reduces abundance |
| *Streptococcus parasanguinis_B* | -27.17 | 6.60e-21 | 7.00e-20 | Reduces abundance |
| *Hungatella hathewayi* | -27.16 | 6.75e-21 | 7.00e-20 | Reduces abundance |
| *Tyzzerella MIC8139* | -26.77 | 2.39e-20 | 2.27e-19 | Reduces abundance |
| *Acutalibacteraceae MIC8041* | -26.64 | 3.66e-20 | 3.28e-19 | Reduces abundance |
| *Blautia hominis* | -26.62 | 3.90e-20 | 3.41e-19 | Reduces abundance |
| *Pauljensenia sp001838165* | -25.89 | 3.98e-19 | 3.00e-18 | Reduces abundance |
| *GCA-900066495 MIC8689* | -25.85 | 4.48e-19 | 3.23e-18 | Reduces abundance |
| *Haemophilus_D sp001679485* | -25.04 | 5.43e-18 | 3.60e-17 | Reduces abundance |
| *Veillonella_A sp000431435* | -24.73 | 1.35e-17 | 8.63e-17 | Reduces abundance |
| *Turicibacter sanguinis* | 23.84 | 1.85e-16 | 1.12e-15 | Increases abundance |
| *Corynebacterium argentoratense* | 23.89 | 1.63e-16 | 1.00e-15 | Increases abundance |
| *Streptococcus sp001587175* | 24.56 | 2.29e-17 | 1.43e-16 | Increases abundance |

| | | | | |
|---|---|---|---|---|
| *QAMM01 sp003150405* | 24.76 | 1.27e-17 | 8.29e-17 | Increases abundance |
| *Dorea sp900066555* | 25.63 | 8.79e-19 | 5.95e-18 | Increases abundance |
| *Clostridium_M lavalense* | 25.75 | 6.16e-19 | 4.26e-18 | Increases abundance |
| *Absiella dolichum* | 25.77 | 5.79e-19 | 4.09e-18 | Increases abundance |
| *Dorea phocaeense* | 25.87 | 4.16e-19 | 3.07e-18 | Increases abundance |
| *CAG-306 sp000980375* | 25.92 | 3.62e-19 | 2.80e-18 | Increases abundance |
| *Clostridium_M MIC9401* | 26.01 | 2.68e-19 | 2.12e-18 | Increases abundance |
| *Klebsiella pneumoniae* | 26.08 | 2.18e-19 | 1.77e-18 | Increases abundance |
| *Holdemania sp900120005* | 26.31 | 1.07e-19 | 8.84e-19 | Increases abundance |
| *UBA11774 sp003507655* | 26.49 | 5.88e-20 | 5.01e-19 | Increases abundance |
| *Massiliomicrobiota sp002160815* | 26.70 | 2.98e-20 | 2.75e-19 | Increases abundance |
| *Streptococcus MIC7545* | 26.90 | 1.58e-20 | 1.54e-19 | Increases abundance |
| *Eubacterium callanderi* | 27.15 | 7.00e-21 | 7.04e-20 | Increases abundance |
| *Holdemanella sp002299315* | 27.22 | 5.59e-21 | 6.19e-20 | Increases abundance |
| *Eubacterium_G sp000432355* | 28.35 | 1.26e-22 | 2.09e-21 | Increases abundance |
| *Catenibacterium sp000437715* | 28.63 | 4.82e-23 | 8.43e-22 | Increases abundance |
| *Faecalibacterium prausnitzii_E* | 28.89 | 1.98e-23 | 3.87e-22 | Increases abundance |
| *CAG-245 sp000435175* | 28.91 | 1.87e-23 | 3.87e-22 | Increases abundance |
| *UBA1691 MIC9213* | 29.14 | 8.20e-24 | 1.94e-22 | Increases abundance |
| *Eubacterium_G sp000435815* | 29.27 | 5.24e-24 | 1.45e-22 | Increases abundance |
| *GCA-900066995 sp900291955* | 29.86 | 6.44e-25 | 2.14e-23 | Increases abundance |
| *CAG-303 sp000437755* | 30.00 | 3.87e-25 | 1.43e-23 | Increases abundance |
| *Bifidobacterium sp002742445* | 30.00 | 3.87e-25 | 1.43e-23 | Increases abundance |

**Supplementary Table 2: Significantly differentially abundant MetaCyc groups pathways (adjusted p<0.05) between the intervention and control groups at end of intervention**

| MetaCyc pathway | Log2FoldChange | p-value | Adjusted p-value |
|---|---|---|---|
| PWY-5297~siroheme amide biosynthesis | 1.0786824 | 0.007677942 | 0.9985634 |
| PWY-6057~dimethyl sulfide degradation III (oxidation) | -0.9713194 | 0.010575704 | 0.9985634 |
| PWY66-426~hydrogen sulfide biosynthesis II (mammalian) | 2.0755318 | 0.001762352 | 0.7798408 |
| PWY-5274~sulfide oxidation II (sulfide dehydrogenase) | 2.0874901 | 0.006540974 | 0.9985634 |
| PWY-5026~indole-3-acetate biosynthesis V (bacteria and fungi) | 1.0169086 | 0.005205804 | 0.9985634 |
| P483-PWY~phosphonoacetate degradation | 1.3121501 | 0.042327007 | 0.9985634 |
| PWY-6826~phosphatidylcholine biosynthesis VI | 1.8847429 | 0.001686858 | 0.7798408 |
| PWY-1263~taurine degradation I | 0.9798989 | 0.031200209 | 0.9985634 |

| | | | |
|---|---|---|---|
| PWY-6510~methanol oxidation to formaldehyde II | 0.7604842 | 0.003013040 | 0.8888469 |
| PWY-6549~L-glutamine biosynthesis III | 4.5490443 | 0.016405530 | 0.9985634 |
| PWY-6348~phosphate acquisition | 0.5002587 | 0.020367565 | 0.9985634 |
| PWY-7761~NAD salvage pathway II (PNC IV cycle) | 1.3745835 | 0.019518295 | 0.9985634 |
